# A mechanistic framework for interpreting blood-stage malaria vaccine efficacy

**DOI:** 10.64898/2026.09.28.26363670

**Authors:** Jason R. Wood, Joseph D. Challenger, Thomas S. Churcher, Lucy C. Okell, Azra C Ghani

## Abstract

Blood-stage vaccines (BSVs) against malaria offer a promising avenue for reducing parasite burden and transmission, yet their efficacy is challenging to interpret across different trial phases. In this study we develop a simple mechanistic within-host mathematical model of *Plasmodium falciparum* asexual parasitaemia. We fit this model to individual-level data from the phase I/IIa RH5.1 trial and estimate the relationship between vaccine efficacy and RH5 antibody titres, enabling the quantitative prediction of vaccine-impact on public health outcomes. We show that small reductions in erythrocyte invasion rate can lead to substantial decreases in probability of exceeding parasitaemia thresholds and transmission potential. Our framework provides a flexible, biologically motivated method for interpreting early-phase trial data and can inform future trial design, vaccine evaluation and integration into broader malaria control strategies.

## Introduction

Malaria remains a serious public health concern. In 2024 there were an estimated 263 million cases and 597 thousand deaths, the majority caused by *Plasmodium falciparum* in Sub-Saharan Africa^1^. Interventions including improved antimalarial treatment, insecticide treated nets and seasonal malarial chemoprevention dramatically reduced the burden of malaria in the late 2000’s, but progress has slowed since 2015^1^.

Recently, two malaria vaccines – RTS,S and R21 - were approved for use by the World Health Organisation (WHO), both of which show high levels of efficacy against clinical malaria caused by *P. falciparum*^2–5^. These vaccines target the pre-erythrocytic stage of the parasite lifecycle (PEVs) before the parasite exits the liver^6^. Whilst highly effective, efficacy is not 100% and hence breakthrough infections occur. In contrast, blood-stage vaccines (BSVs) target the asexual parasites which develop after being released from the liver. One of the most promising current candidates, RH5, works by stimulating production of antibodies and other immune responses which bind to surface proteins on the merozoite, preventing the invasion of erythrocytes^7^. Other blood-stage vaccine candidates such as PfGLURP target the trophozoite and schizont stages by triggering immune responses against the parasite proteins on the surface of infected erythrocyte, causing it to be broken down before the release of the next generation of merozoites^8^. Both targets work to reduce the number of parasites produced in the subsequent generation of asexual parasites, reducing the observed parasite multiplication rate (PMR).

Early human testing of malaria vaccines occurs using controlled human malaria infection (CHMI) trials, whereby participants are challenged either through infection with sporozoites or blood-stage parasites^9,10^. In these trials, for PEVs, efficacy is defined as the relative risk of blood-stage infection following either infectious bite from a mosquito or inoculation with sporozoites in the vaccinated population compared to the risk of infection in the control population^11^. However, BSV trial efficacy is instead defined on the basis of the reduction in parasite multiplication rate (PMR)^12^. PMR is obtained by calculating the log-linear growth of parasite density, and the reduction in PMR caused by the vaccine is then calculated by comparing the PMRs for each vaccinated individual with the mean PMR of the control populations^12–14^.

Translating efficacy from CHMI trials of BSVs into classical efficacy measures expected in field trials (i.e. reduction in clinical symptomatic malaria incidence^15^) is not straightforward. For example, in a CHMI trial, the current candidate RH5.1 showed ∼20% efficacy in reducing PMR in vaccinated individuals^12^ whereas in the subsequent phase II field trial, it demonstrated a ∼55% efficacy against malaria incidence (subject to threshold of parasitaemia)^15^. Without an understanding of how reductions in PMR translate into reductions in infection/disease, it is dificult to prioritize next generation candidates for field study.

Here we developed a mechanistic within-host mathematical model to estimate the efficacy of BSVs from CHMI trial data, showing that estimates of the reduction in erythrocyte invasion rate correlate closely with the reduction in PMR. Our results demonstrate that small reductions in erythrocyte invasion rate can significantly decrease both parasitaemia thresholds and onward transmission potential. Our modelling framework offers a biologically motivated method to interpret early-phase trial data, supporting the design and evaluation of future blood-stage malaria vaccines.

## Results

### Dynamics of asexual parasitaemia

We first developed a mechanistic mathematical model to capture the within-host dynamics of an early-stage asexual, blood-stage *P. falciparum* infection following blood-stage challenge (Model 1). To do so, we extended a previously developed model^16^ to capture the mechanism of the RH5.1 vaccine which acts to reduce the rate at which parasites invade red blood cells (i.e. the erythrocyte invasion rate) (Figure 1a). We fitted this model to the parasitaemia measurements from the CHMI challenge as part of a phase I/IIa trial of RH5.1 recombinant protein vaccine^12^ (Figure 1b).

**Figure 1:**
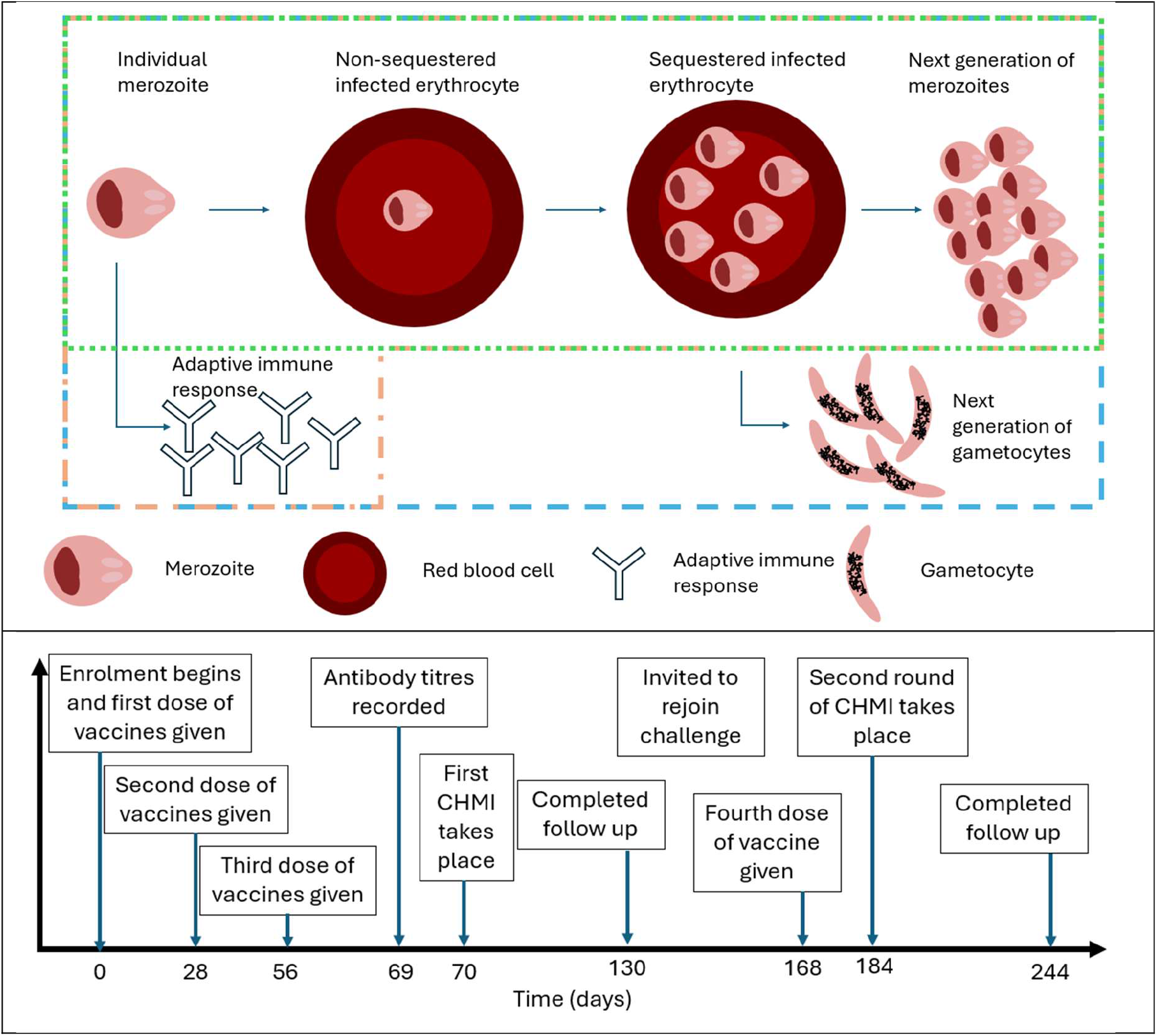
(A) Biological depictions of the three models described mathematically in the Methods section, Model 1 considers only the dynamics of the parasites and is captured by the green box. Model 2 introduces an adaptive immune response and is contained in the orange shape. Model 3 also introduces gametocyte dynamics and is contained in the blue box. (B) Timeline of the RH5.1 CHMI trial conducted in Oxford, generated using the information available in Minassian et al. (2021)^12^.

The fitted model was able to capture the typical dynamics observed within the early stages of a blood-stage malaria infection for both unvaccinated and vaccinated individuals (Figure 2). Model fits for all participants using the median posterior estimate of participant-specific draws of the erythrocyte invasion rates are shown in Figure S9 (Supplementary Material).

**Figure 2:**
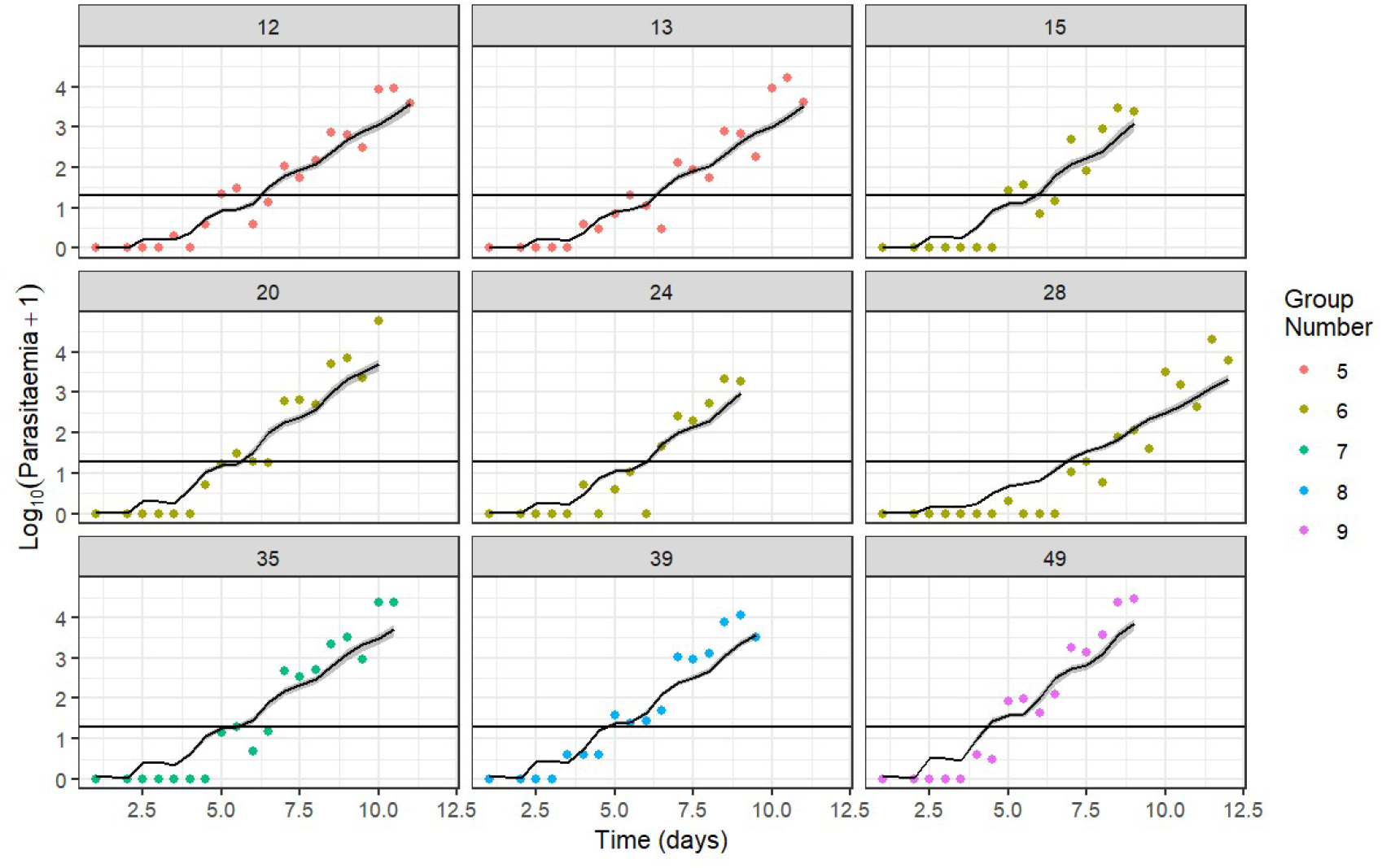
Model projected growth of blood-stage asexual parasitaemia of a random selection of individuals from the RH5 trial measured by qPCR following challenge at day 0. The central lines show the median modelled estimate using samples from the posterior distribution calculated for the individuals. Different dot colours denote the different groups from the challenge. Shaded regions represent 95% credible interval. Parameters as in Table 2 except for sampled parameters. Horizontal black lines denote 20 parasites/mL, the limit of detection.

### Estimation of RH5 vaccine impact on parasitaemia

We estimated the maximum vaccine efficacy against erythrocyte invasion rate, *V*_*max*_, to be 35% (95% CrI – 14%-55%), with median trial efficacy of 28.6%. Estimated efficacy increased with antibody titre (Figure 3A), with 50% of the efficacy achieved at a titre value of 6.67 (95 % CrI – 0.18-71.7) anti-RH5_FL IgG (*µg*/*mL*) and 90% of the maximum efficacy achieved at a titre value of 61.0 (95 % CrI – 20.5 -652.5) anti-RH5_FL IgG (*µg*/*mL*). When estimating these parameters, we excluded two outlier individuals. The effects of including these individuals as well as a sensitivity analysis to the fixed parameters in Table 1 are shown in Figures S1-8.

**Table 1:**
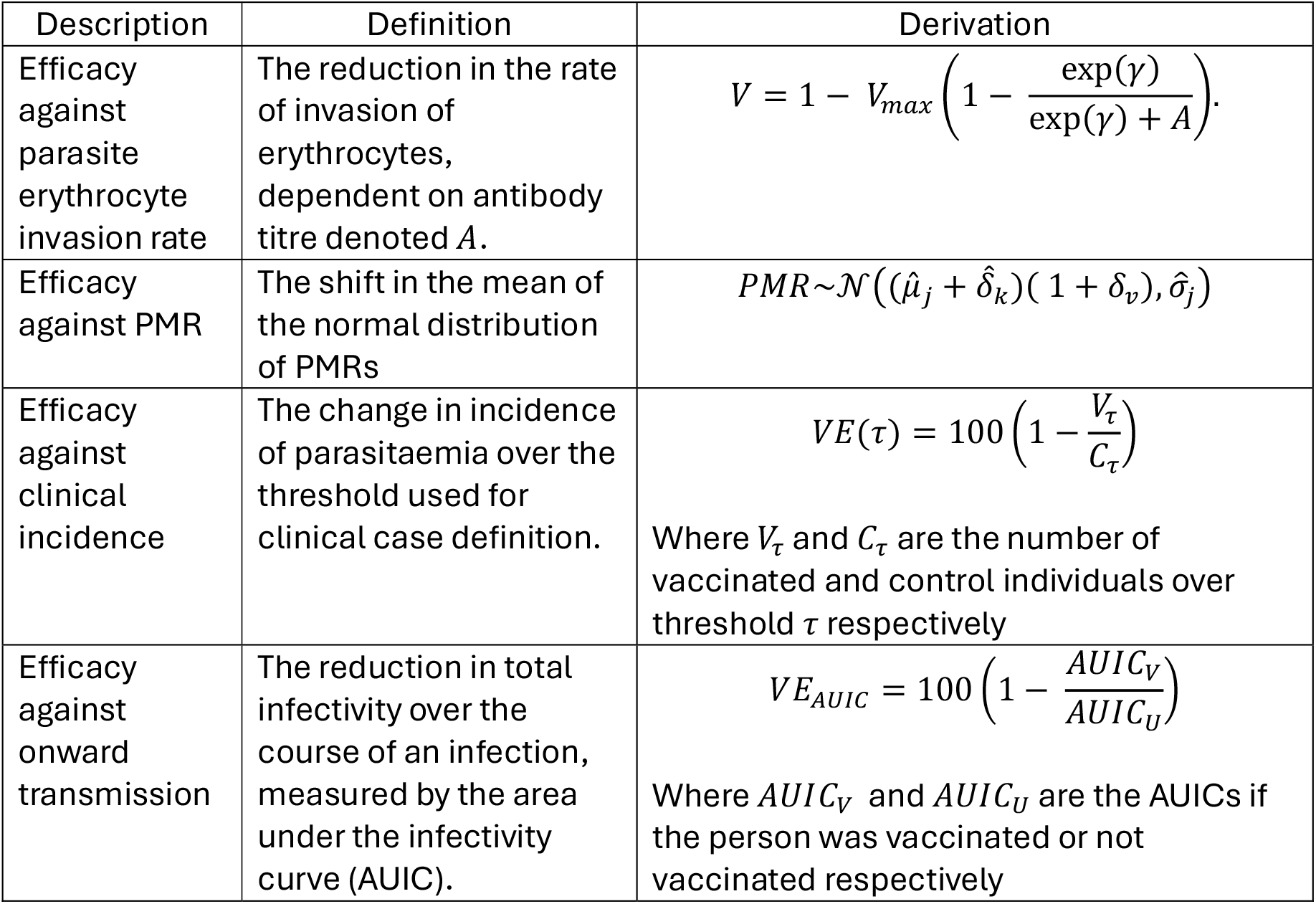
Descriptions, definitions and calculations of different forms of vaccine efficacy.

**Table 2:**
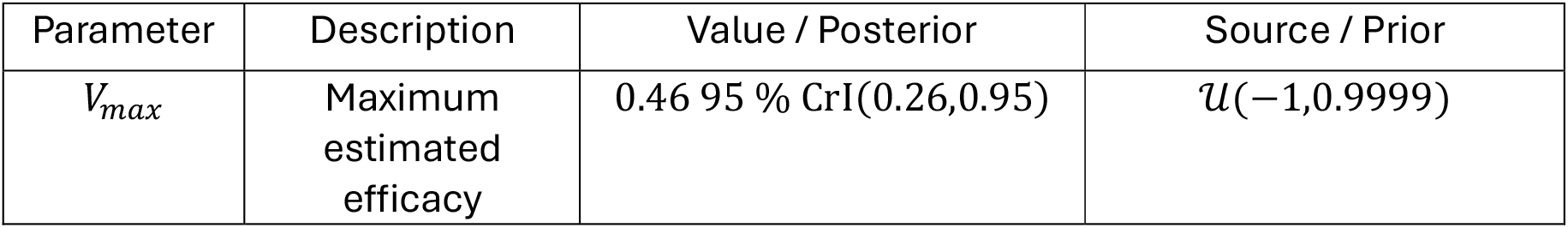
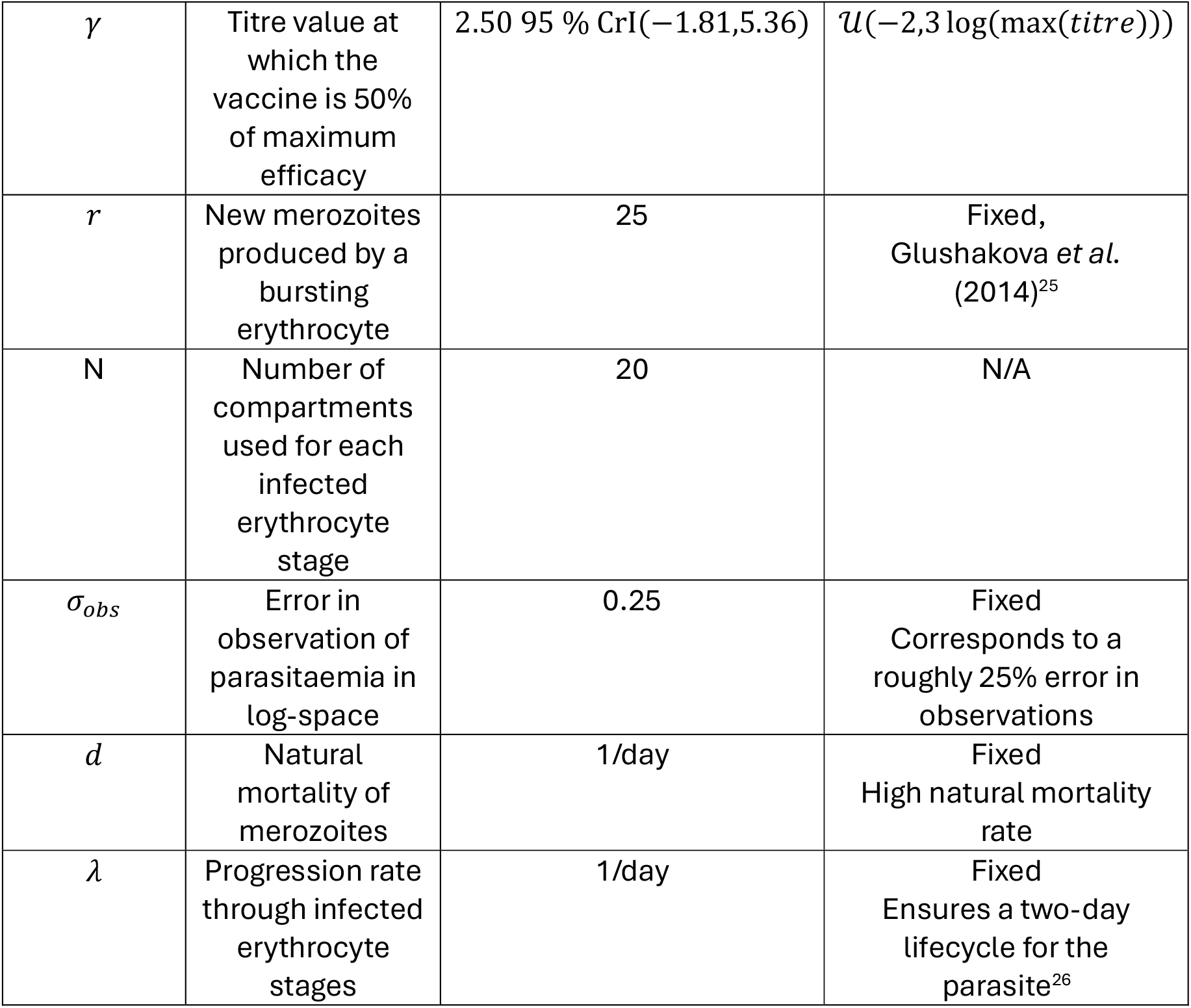
Parameters of the model described in Equations 1-5. Additionally, posteriors and priors for the estimated parameters of the BSV RH5.1. For the two fitted parameters, the table shows the prior and posterior estimates from fitting to the trial data.

| Parameter | Description | Value / Posterior | Source / Prior |
| --- | --- | --- | --- |
| $V_{max}$ | Maximum estimated efficacy | 0.46 95 % CrI(0.26,0.95) | $\mathcal{U}(-1, 0.9999)$ |
| $\gamma$ | Titre value at which the vaccine is 50% of maximum efficacy | 2.50 95 % CrI(−1.81,5.36) | $\mathcal{U}(-2,3 \log(\max(\text{titre})))$ |
| $r$ | New merozoites produced by a bursting erythrocyte | 25 | Fixed,<br>Glushakova *et al.* (2014)<sup>25</sup> |
| N | Number of compartments used for each infected erythrocyte stage | 20 | N/A |
| $\sigma_{obs}$ | Error in observation of parasitaemia in log-space | 0.25 | Fixed<br>Corresponds to a roughly 25% error in observations |
| $d$ | Natural mortality of merozoites | 1/day | Fixed<br>High natural mortality rate |
| $\lambda$ | Progression rate through infected erythrocyte stages | 1/day | Fixed<br>Ensures a two-day lifecycle for the parasite<sup>26</sup> |

**Figure 3:**
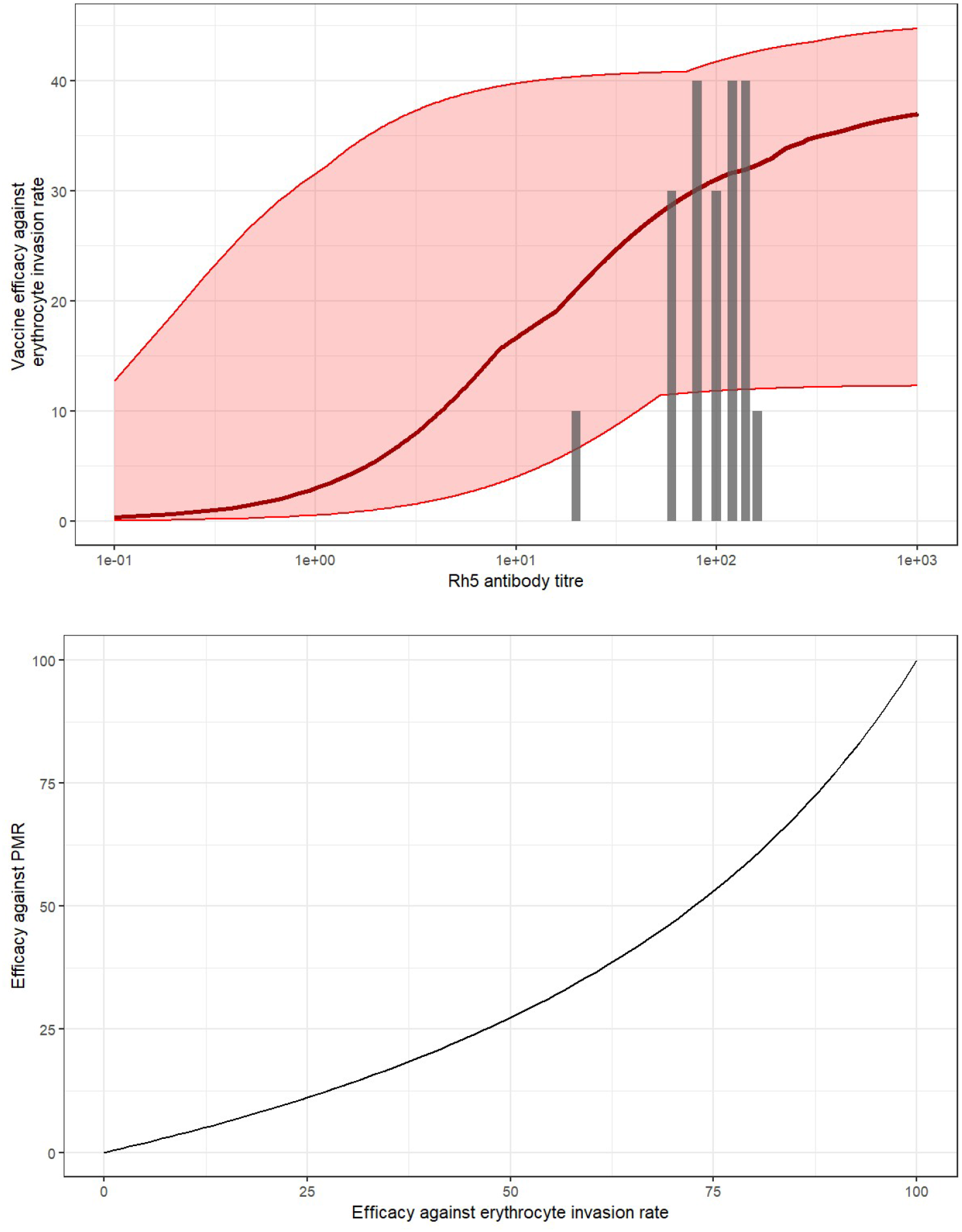
(A) Relationship between vaccine efficacy against erythrocyte invasion rate and anti-RH5_FL IgG titre (µg/mL). The red line shows the median relationship between antibody titre and vaccine response, and the shaded region shows the 95% credible interval. Vertical grey bars show the distribution of the antibody titres from the challenge study participants. (B) A demonstration of the relationship between the efficacy against erythrocyte invasion rate and the efficacy against PMR

In CHMI studies for blood-stage vaccines, the reduction in the parasite multiplication rate (PMR) is used as a measure of vaccine efficacy. This is related to our biologically motivated estimate of the efficacy against the erythrocyte invasion rate as shown in Table 1 and Figure 3B. To assess the ability of our model to act as a potential trial simulator relating these metrics, we simulated a synthetic cohort of trial participants whose antibody titre levels and parasite erythrocyte invasion rates were generated using Model 1 using our fitted parameter estimates. Our estimate of the reduction in PMR from the synthetic cohort (21.7%, 95% CrI, 14.0%-29.9%) is comparable to the data estimated reductions in PMR in the phase 1 trial (∼20%).

### Efficacy against clinical incidence

Whilst the effect of blood-stage vaccines is a reduction in the growth rate of asexual parasitaemia, this is not observable in field trials, where efficacy focuses on comparison of clinical incidence between vaccinated and control arms^15^. As definitions of clinical disease include threshold parasitaemia levels, we used our model to explore the relationship between our biological vaccine efficacy measure and vaccine efficacy against clinical disease (Table 1).

Figure 4A illustrates how the threshold parasitaemia used to define clinical incidence influences vaccine efficacy against clinical incidence. For low thresholds (e.g. 1,000 parasites/mL shown in red), there is no discernible difference in clinical incidence rates between the unvaccinated and vaccinated cohorts, except at unrealistically high (*V*_*max*_ ≥ 0.95) levels of vaccine efficacy against erythrocyte invasion. However, at more commonly used thresholds (e.g. 5,000,000 parasites/mL shown in purple), we observe a near-linear relationship between vaccine efficacy against erythrocyte invasion and vaccine efficacy against clinical incidence. For the RH5.1 CHMI data we obtained an estimate of maximum vaccine efficacy against erythrocyte invasion of 35%. This translates to an estimated vaccine efficacy against clinical incidence of 37.1% (95% CrI 12.9-56.2).

**Figure 4:**
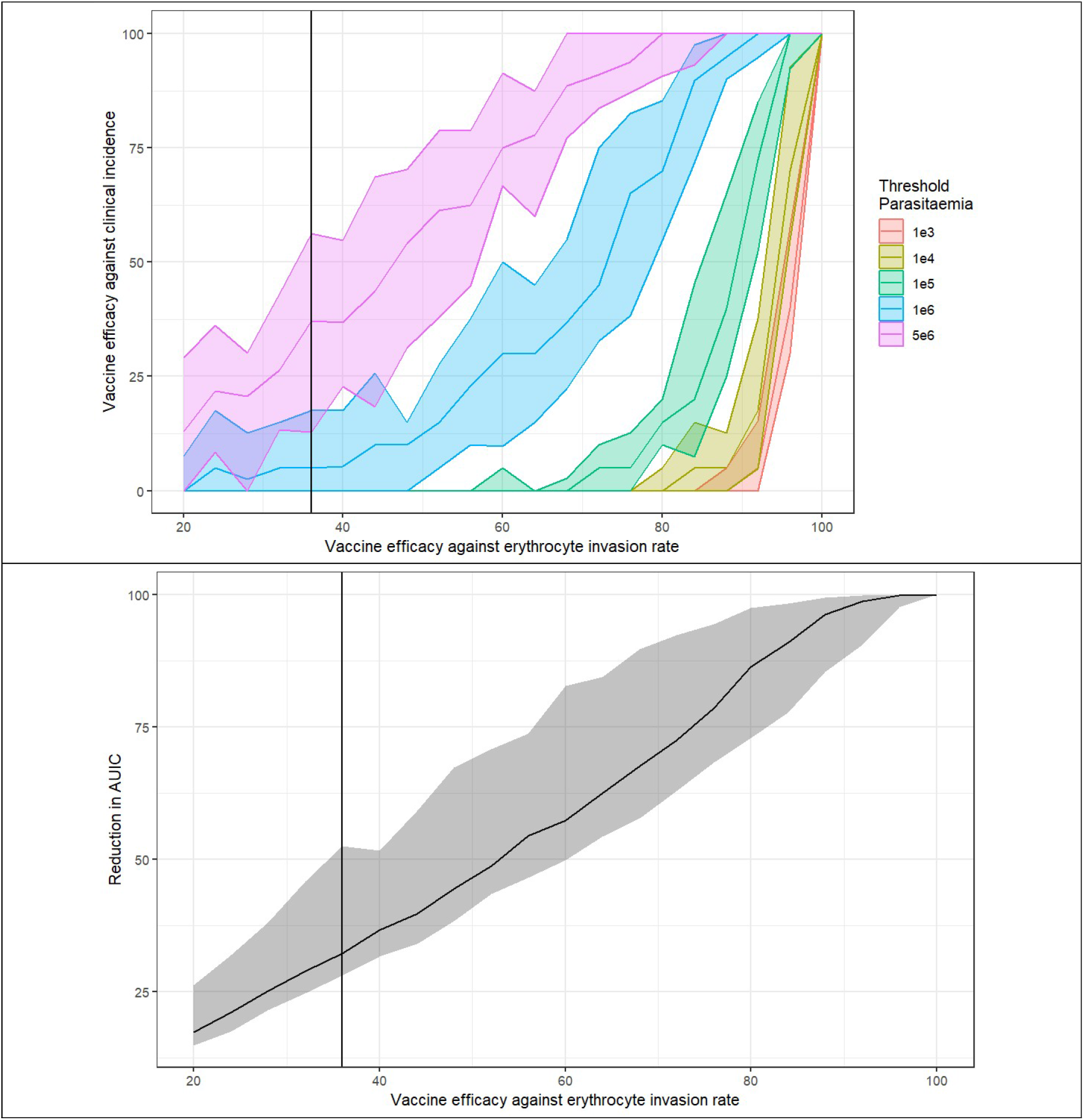
(A) Vaccine efficacy against clinical incidence plotted against vaccine efficacy against erythrocyte invasion, a vertical black line shows the median efficacy of RH5. Vaccine efficacy against clinical incidence is defined as the reduction in the probability of exceeding the threshold given the individual received the vaccine (Table 3 for details). Different thresholds are denoted in different colours. Credible intervals are calculated using repeated sub-sampling. Example longer term simulations are available in Figure S11. (B) Reduction in Area under the infectivity curve (AUIC) for increasing vaccine efficacies. The solid black line depicts the median reduction due to the vaccine, and the shaded region shows the 95% credible interval. The vertical line denotes a vaccine as effective as RH5. Credible intervals calculated using a bootstrapping scheme.

### Blood-stage vaccines reduce transmission potential

By reducing circulating parasitaemia, blood stage vaccines could also reduce onward transmission by reducing the production of gametocytes. To explore this, we extended the model to capture gametocyte production (Equation 3) and incorporated a previously derived statistical relationship between gametocyte density and infectivity to mosquitoes. We defined vaccine efficacy against onward transmission as the reduction in the area under the infectivity curve as previously described by Bradley et al.^17^.

Figure 4B shows the simulated relationship between vaccine efficacy against erythrocyte invasion and vaccine efficacy against infectivity. We observe a near-linear relationship between these two measures of vaccine efficacy, suggesting strong potential for effective blood-stage vaccines to also impact onward transmission.

## Discussion

We developed a mathematical model of the blood-stage asexual dynamics during the early stages of a *P. falciparum* infection in malaria naïve adults. We have fit this model to individual-level parasitaemia data from the phase I/IIa vaccine trial for RH5^12^ to translate reductions in erythrocyte invasion rate into reductions in clinical malaria incidence. This modelling framework can be used to fit to data from other blood-stage vaccines, and it is possible to use it in both an antibody-dependent and to simply directly model effects on erythrocyte invasion rate, allowing for direct comparisons between blood-stage vaccine candidates (see Supplementary Material for a fitting to the AMA1 CHMI data).

Our results highlight the differences between measures of efficacy in phase 1 and phase 2 trials, and how that may in turn lead to different assessments of vaccine efficacy. Due to the non-linear relationship between parasite replication, parasite erythrocyte invasion rate, and parasitaemia thresholds required to meet case definitions of clinical malaria, a vaccine which reduces the parasite erythrocyte invasion rate by 50% may only reduce the probability of exceeding the pyrogenic threshold by 30-40%. This figure also highlights how the choice of clinical threshold can influence observed efficacy. Increasing the parasitaemia threshold increases the observed efficacy, though this effect vanishes at very high levels of vaccine efficacy against erythrocyte invasion rates. This corroborates the results seen in the interim results of the phase 2b trial for RH5 in Burkina Faso, where the efficacy of the vaccine depended on the threshold selected, varying from 15% efficacy at the lowest threshold to 91% efficacy at the highest threshold^15^.

From the interim phase 2b trial results, the efficacy of 55% or 40% depending on whether the cohort was vaccinated in a delayed or monthly scheme respectively. This 40% efficacy is similar to our estimates of efficacy when using the same threshold, 5,000,000 parasites per millilitre is 5,000 parasites per microlitre, the threshold used in the phase 2b trial. Despite mathematical models never capturing all the details available, our analysis has done a good job of translating phase 1 efficacy into sensible phase 2 results.

The non-linear relationship between efficacy against erythrocyte invasion rate and efficacy against clinical infection is important as large population sizes may be needed in phase 2 trials and beyond to see the predicted effects. For instance, using the threshold of 100,000 (1e5) parasites/mL in Figure 4A, we can see that for two vaccines, one with 40% efficacy against erythrocyte invasion rate and one with 80% efficacy against erythrocyte invasion rate, the increase in efficacy against clinical incidence would be around 12%. Whereas if we selected the 1,000,000 (1e6) parasites/mL threshold in Figure 4A, to give a ∼60% increase efficacy. This significant difference in expected efficacy would make it possible to power clinical trials to detect the effect with significantly fewer participants, as standard power calculations use relative rates of events^18^.

When exploring the longer-term consequences of blood-stage vaccination and increasingly efficacious vaccines, we utilised a simplified model of the immune dynamics. This simplified model neglects some of the important aspects of the interactions between the *P. falciparum* parasite and the host immune system, such as var-gene switching or the decay of vaccine induced antibodies. Neglecting these effects does not meaningfully change our interpretation, as the main purpose of the onwards analysis was to demonstrate the effects blood-stage vaccines may have on levels of parasitaemia reached and onwards transmission.

Our analysis also suggested that blood-stage vaccines could have significant benefit in reducing onwards transmission. Vaccinated individuals had lower gametocyte AUICs than non-vaccinated individuals, with at least a 15% reduction in AUIC at the lowest level of vaccine efficacy that we explored. Reductions in AUIC do not necessarily correspond to an equivalent reduction in the number of mosquitoes infected, as this will depend on the frequency of bites and the probability of transmission when bitten. Due to the BSV, the timing of the peak of parasitaemia – which will likely be lower - is increased. This delay may flatten the peak of malaria incidence at a population level, potentially reducing onwards transmission during high-risk periods. The impact of blood-stage vaccines on onwards transmission is predicted to be substantial and could have important epidemiological impacts. Measuring human-mosquito transmission though direct membrane feeding assays is logistically challenging and expensive so methods such as these, which infer these population-level impacts, may have utility, though further work is needed to verify assumptions used in the model Overall, our modelling framework offers a tool for interpreting early phase clinical trial data and translating to outcomes of reductions in clinical malaria at the population-level. This framework could be used to inform trial design, helping to identify antibody thresholds which have important implications for onwards transmission or symptomatic disease. This work can also be taken further through combination with transmission models to design public health interventions. Further work should also consider possible synergistic effects that might emerge from the combination of multiple interventions, for instance the co-administration of PEV and BSV vaccines.

## Methods

### Trial data

We use publicly available data from a RH5.1 phase IIa CHMI trial conducted in Oxford, UK^12^ (Figure 1b). 16 males and 22 females were enrolled, with a mean age of 27 (range 21-39). 31 participants were recruited in the first round of the trial, and 9 additional participants were recruited for the second round of challenges. The initial 31 participants were divided into 2 initial arms (vaccination & control). A subset of the 31 participants undertook a subsequent re-challenge, with the vaccinated re-challengees receiving a booster dose. The 9 additional participants recruited into this secondary round served as a re-challenge control arm.

For those in the vaccine arm, the primary 3-dose vaccine schedule was administered on day 0, day 28 and day 56. Participants (including the control arm) were challenged 14 days after dose 3 (day 70, denoted dC). Follow up continued for 90 days after challenge.

For the rechallenge subgroup, for the vaccine arm a fourth vaccine dose was administered on day 168. The subset were then rechallenged on day 183 (dC^2^) and the subgroup was followed for a further 90 days.

There were 52 challenges in total, across 35 unique individuals, with 17 individuals challenged twice. Challenge participants were inoculated with a dose of ∼1000 erythrocytes infected with 3D7 *P*.*falciparum*.

Antibody titres were measured one day before challenge (d69 and d182) and parasitaemia was subsequently recorded by qPCR twice a day. Parasitaemia was recorded in this way until treated according to a predefined diagnostic algorithm, except for one participant in the second round who withdrew from the trial. Further information is available in the supplementary material of Minassian *et al*. (2021)^12^.

### Within-host model of blood-stage dynamics

We sequentially extended a set of deterministic models to capture the blood-stage dynamics of the different phases of a *P. falciparum* infection, building on the framework presented in Khoury *et al*. (2018)^16^. The dynamics of asexual parasitaemia during *P. falciparum* infection are often described using discrete time models^19–21^ with two-day timesteps to account for the lifecycle of the parasite. However, owing to the frequency of sampling in the RH5.1 trial (see later section), we opted to use a continuous time model. This model construction also allows us to more naturally specify mechanism of action of the RH5.1 vaccine (i.e. blocking infection of erythrocytes).

In all models, *P* is the density of merozoites per millilitre (mL), *S* is the density of the detectable, non-sequestered infected erythrocytes, and *I* is the density of the undetectable, sequestered infected erythrocytes per mL be denoted *I*. Merozoites infect uninfected red blood cells at a rate *β* and die due to natural mortality and innate immune response mechanisms (e.g. splenic clearance^22^) with rate *d*. Infected erythrocytes burst after progressing through *2N* stages with rate *λ*, where *N* is the number of compartments. As the number of compartments *N* increases, we shift the time spent in the erythrocyte from being exponentially distributed with mean 2 days, to being Erlang distributed with mean 2 days and shape *N*. When infected erythrocytes exit the final compartment they burst, producing *r* new merozoites with rate *λN*. For the first *N* stages, the parasites are detectable, after which they are assumed to sequester. This is equivalent to assuming that parasites on average spend one day sequestered. All values of fixed parameters are in Table 2.

### Model 1

Model 1 is used to estimate the effect of the blood-stage vaccine on parasite growth, using the data from the RH5.1 phase IIa trial^12^. As all trial participants were treated within 14 days (except one trial participant in the control rechallenge group who displayed no noticeable parasite growth over 20 days) the uninfected red blood cell population is assumed to be constant. Additionally, as it is well known that other naturally acquired immune responses to blood-stage malaria develop slowly, we assume that no adaptive immune response is induced by the blood-stage dynamics during the trial. We further assume that the immune response induced by the vaccine is constant. This is likely to be an oversimplification, as from Minassian *et al*. (2021)^12^ we know that the antibodies do decay over time, but due to the short length of the challenge this is unlikely to have a substantial effect. Finally we assume that the inoculum in the CHMI is consistent within a round of challenges, but that there is twice the inoculum in the second round of challenges, owing to the reported size of inoculation^23^.

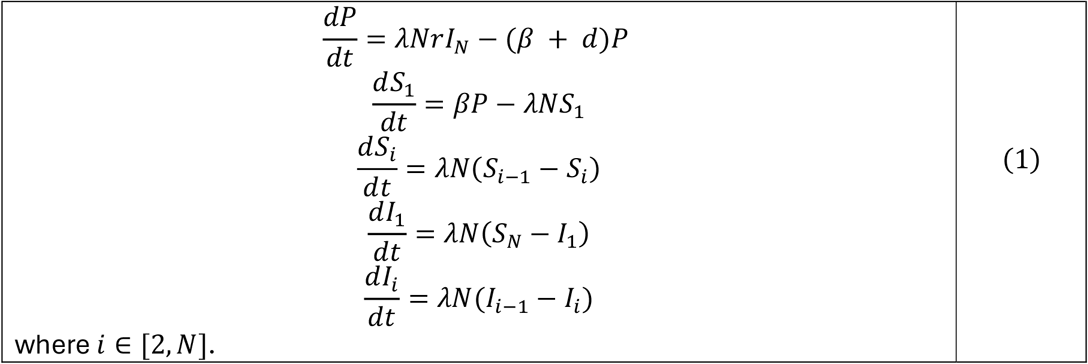

### Model 2

Model 1 does not capture clearance of infection. We extended the model to allow for the effect of a vaccine independent adaptive immune response developed through a density-dependent process (Equation 2). This imposes an upper limit to parasitaemia, after which parasite density will decay to zero. We assume that only merozoites invoke the adaptive immune response, denoted by *A*, with rate *ω*, and that the immune response clears merozoites with rate *ϵA*. Here we assume that all individuals are malaria naïve.

We use this model to estimate the impact of the vaccine on clinical incidence defined by parasite density.

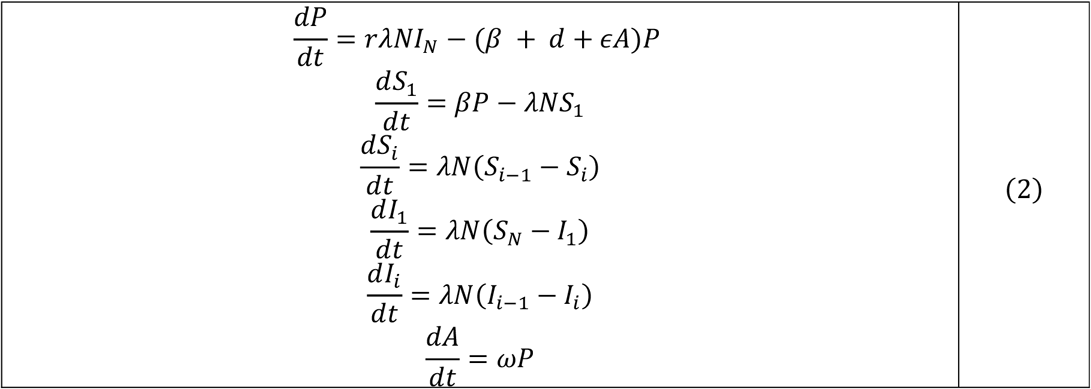

### Model 3

The effects of the reduced parasite growth on the production of gametocytes may also be an important component of the public health impact of a BSV. Here we extend Model 2 with a minor modification of a previously developed mathematical model for gametocytaemia by Challenger *et al*. (2019)^19^. We modify this model by allowing direct progression from the infected erythrocytes into immature gametocytes. Here gametocytes mature through five stages, with three compartments per life stage, and are denoted by 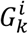, with *i* denoting the compartment and *k* denoting the life stage.

Merozoites sexually commit with rate *α* and the progression rate through the life stages is denoted by *δ*_*k*_ and is the same for the first four life stages and differs in the fifth stage. This model structure and the parameters used result in gametocytaemia lagging parasitaemia by around 15 days.

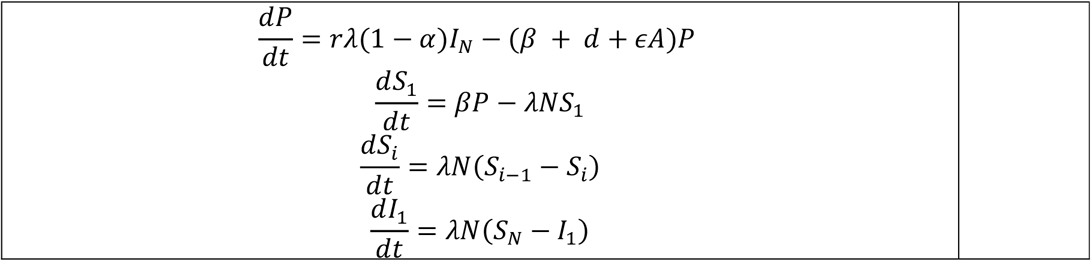

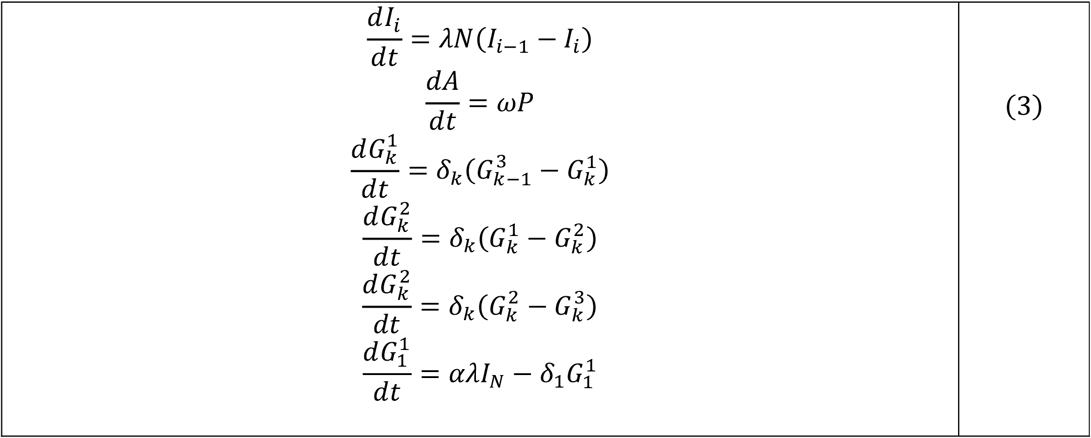

Here *δ* = (*kL, kL, kL, kL*, k_5_*L*), and *L* = 3 is the number of auxiliary compartments used in the gametocyte model. In the work of Challenger *et al*. (2019)^19^ the parameters *α, k* and *k*_5_ are distributed according to a truncated Normal distribution. Here we use the means 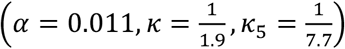 of these distributions.

### Measures of vaccine efficacy

#### Efficacy against erythrocyte invasion rate

We consider a model where vaccine efficacy depends on antibody titres. We model the effects of the vaccine-induced antibodies using an exponential formulation of the Hill function, similar to the formulation used for the modelling of the pre-erythrocytic vaccines RTS,S and R21^5^. The equation is,

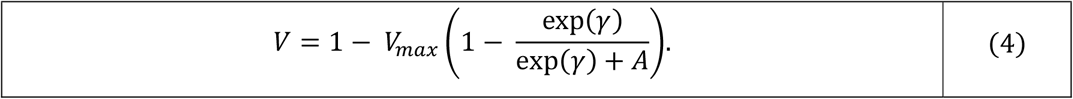

where *A* is the antibody titre of interest,*γ* is the log antibody titre value at which the vaccine causes a reduction in the erythrocyte invasion rate of the merozoites by 50% of the maximum possible efficacy, and *V*_*max*_ is the maximum possible efficacy of the vaccine. We assume that the presence of the vaccine modifies an individual’s erythrocyte invasion rate *β*, such that,

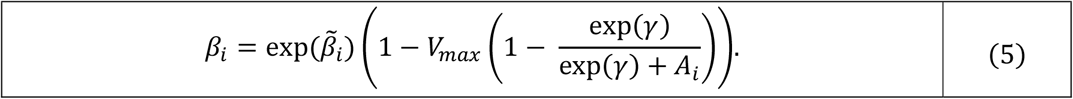

where *A*_*i*_ is the individual’s antibody titre, and 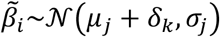, where *µ*_*j*_ is the global mean of the erythrocyte invasion rate for trial round *j, δ*_*k*_ is the shift in the mean of the erythrocyte invasion rate caused by being previously challenged, subscript *k* denotes whether an individual was previously challenged, and *σ*is the standard deviation in the log of the erythrocyte invasion rates for trial round *j*. When *A*_*i*_ = 0, the expression in equation 5 reduces to 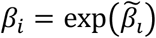.

The models were fitted to the trial data using Markov Chain Monte Carlo (MCMC) implemented using the adaptive random walk algorithm from the monty package (version 0.3.24) in R^24^. In all instances, 4 chains were used, with 20000 sampling steps with a 10000-step burn-in period. The observations of parasitaemia were assumed to have a log-Normally distributed error when above a threshold, and a censored observation likelihood otherwise. When the observed parasitaemia *x*_*i*_ is above the threshold the likelihood is

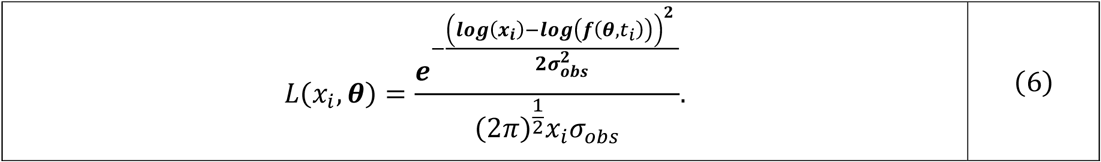

When the parasitaemia *x*_*i*_ is below the observation threshold, the likelihood is given by

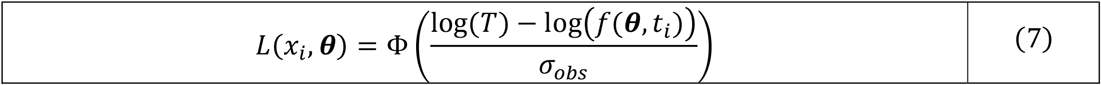

where Φ is the cumulative distribution function of the Normal distribution and *x*_*i*_ is the observation value, *T* is the threshold of parasitaemia and *θ* is the parameter vector. The observation function *f* (*θ, t*_*i*_) sums the density of all non-sequestered parasites *S*_*j*_ at Time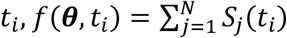.

We removed two outlier individuals from the rechallenge control group (Group 8) from the analysis, as one never experienced any parasite growth, and the other experienced a significant reduction in parasite growth that was not observed when challenged a third time^23^. In the supplementary material, we explore the consequences of not removing these two individuals.

Here we are determining *P*(*V*_*max*_,*γ, µ, σ, δ, β* |*x*). For the parameters *V*_*max*_ and*γ* we assumed uniform priors bounded by [−1,0.9999] and [−2,3 log(*max*(*titre*))] respectively, and for all other fitted parameters we assumed completely uninformative priors. The value of *V*_*max*_ = 1 is excluded due to creating divide by zero errors when rearranging Equation 5.

#### Relationship between efficacy against PMR and efficacy against erythrocyte invasion rate

In our model, the relationship between *β* and the PMR is given by:

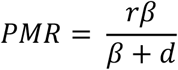

Thus, a reduction *δ*_*p*_ in the PMR is related to a reduction *δ* in *β* by:

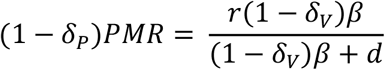

Rearranging this expression gives the relationship between VE measured by PMR and VE measured by the erythrocyte invasion rate:

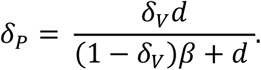

When *δ* = 0 or *δ*_*v*_ = 1 then *δ*_*p*_ = 0 and *δ*_*p*_ = 1, however for all other values this relationship is sublinear, meaning that at any point the curve created by the above equation is beneath *δ*_*p*_ = *δ*_*v*_. The extent of this sub linearity is dependent on the relative size of *β* and *d*, with higher erythrocyte invasion rates leading to slower increases in *δ*_*p*_.

#### Calculating PMR

We validate our synthetic cohort by estimating the vaccine efficacy against PMR and confirming that the efficacy in the synthetic cohort is similar to the efficacy in the phase IIa trial. We use the model of parasitaemia using a PMR structure that is used in Minassian *et al*. (2021)^12^ is

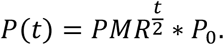

Where *P*_O_ is the initial parasite density, *PMR* is the participants individual parasite multiplication rate, and *t* is the time in days since the inoculation. In the same way we estimate individual erythrocyte attack rates, we estimate individual *PMR*s which are drawn from normal distributions, 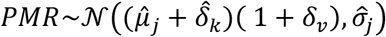. Where similarly to before, 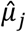 is the mean PMR in trial round j, 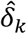 is the effect of one previous challenge, 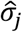 is the standard deviation in trial round j. Here we instead consider the effect of the vaccine to shift the mean of the distribution by *δ*_*V*_.

#### Efficacy against clinical incidence

Febrile malaria is typically associated with parasitaemia exceeding 1,000 parasites/*µ*L or 1,000,000 parasites/mL, though this varies with season and pre-existing immunity^27^. By sampling a synthetic cohort (see Supplementary Material for details), we explored how effective the vaccine is against threshold densities of parasitaemia. The effect of the vaccine is assumed to be constant over the length of the infection as we currently have limited information on the decay rate of the vaccine induced antibodies. We ran model 2 for 100 days, ensuring all infections reach their peak which typically occurs within 18 days. For each erythrocyte invasion rate drawn we performed two simulations with and without vaccination and recorded the maximum parasitaemia level. We used this to calculate the probability of exceeding different parasitaemia thresholds stratified by vaccine status to calculate vaccine efficacy against clinical incidence.

#### Efficacy against onward transmission

To calculate the onwards transmission of an infection, we need to map gametocytaemia to probability of infecting a mosquito. Here we use the statistical relationship developed in Bradley *et al*. (2018)^17^. From Bradley *et al*.^17^ the proportion of mosquitos developing oocysts following biting someone with gametocyte density *x* is,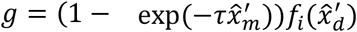. Here 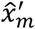 is the density of male gametocytes and 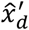 is the density of the population of interest. The statistical relationship with the lowest DIC, used both the male and female density of gametocytes. As our model is theoretical, we do not have a ratio of male-female gametocytes, we therefore use a ratio of 1:4 male: female gametocytes, taken from the median of Figure 1C in Bradley *et al*. (2018). The functional form of 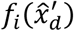 which had the lowest DIC has the form,

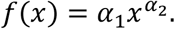

When employing this relationship, we use the median estimated values (τ = 0.08573, *α*_1_ = 0.07108, *α*_2_ = 0.3021) of the parameters as our focus is on the trend produced by the effect of the vaccine rather than propagating uncertainty.

Through these models we can calculate the probability that a mosquito becomes infected given it bites at a specific time point. As in Challenger *et al*. (2019) we then calculate the area under the infectivity curve (AUIC) to allow for a meaningful definition for the effect of the vaccine on onward tranmission^19^.

As when modelling the probability of exceeding a given threshold, we generate a synthetic cohort using the parameters estimated using the RH5 trial data (Supplementary Material). AUIC reduction is calculated on an individual-level basis, comparing the area under the curve if the individual was vaccinated against the area under the curve if they were not. Here we assume that vaccination with a BSV does not affect the rate of commitment to gametocytogenesis, as this is currently unknown^28^.

## Data Availability

All data used in the study are available online.
The data used for the RH5 analysis was located in the supplementary material of the article "Reduced blood-stage malaria growth and immune correlates in humans following RH5 vaccination" and is available at this link https://www.cell.com/cms/10.1016/j.medj.2021.03.014/attachment/3449fc07-92c4-4e2f-9a17-783b6304e570/mmc2.zip
The data for the AMA1 trial was taken from the supplementary material of "Demonstration of the Blood-Stage Plasmodium falciparum Controlled Human Malaria Infection Model to Assess Efficacy of the P. falciparum Apical Membrane Antigen 1 Vaccine, FMP2.1/AS01", available at this link https://oup.silverchair-cdn.com/oup/backfile/Content_public/Journal/jid/213/11/10.1093_infdis_jiw039/2/jiw039_Supplementary_Data.zip?Expires=1793355546&Signature=E4jx7qzqxT4xyaWRdJt7kArXK514224G~PdljhmQ4TzA5V5d--oIYJ1ja8zU2mmPwsMOEmYerVQQuVqVKD0d5P1p1Dr2dk2B6GnYHravDsDL18udtKKHGZ17gffsq-laYymqAo1mpXYgM6yuGNkb1h2Jz1Ji3CK1PQH8eA35Dhs2JqwFzHu6zqn308xtMBNM76kwlt4gGPmPqk2tz-nQrZ8t-F-yx4CtneyvwqhsLAlGr2ul2GH-DjhAWdjVeHUUoVxsDFi7hHipHOcenAIZ0FPD1hWroTZKXj4TY6Y2e8yC1VD0tNykCBJzLGC92C6wZwc4Sv-XGaOpnUNEOUi7GA__&Key-Pair-Id=APKAIE5G5CRDK6RD3PGA

## Ethical approval

All work performed in this study is secondary data analysis of publicly available data. The data was sourced from trial (NCT02044198) and trial (NCT02927145), both of which received appropriate ethical approval.

## Acknowledgements

This work was supported by a joint investigator award to ACG and Prof Katharina Hauck from the Wellcome Trust (220900/Z/20/Z).

