## Supplemental material for "A mechanistic framework for interpreting blood-stage malaria vaccine efficacy"

### Supplementary Materials

### Fitting to AMA1 vaccine CHMI trial data

Using a very similar scheme to the one we used to fit the RH5 data (see Methods), we analyse the effect of AMA1 vaccine using the data generated by the controlled human malaria infection model[1]. The data for the AMA1 vaccine comes from a study demonstrating the ability to use CHMIs to assess the efficacy of BSVs. Participants were inoculated with 1000 (intended size) 3D7 clone-infected erythrocytes, and then parasitaemia was assessed once on day of challenge + 1 and twice a day afterwards using qPCR. 45 volunteers were recruited and split across 3 trial sites, however to our knowledge only the blood-stage parasitaemia for 27 of the participants is reported. The blood-stage parasitaemia was reported for 12 vaccinees and 15 control participants. As we did not have access to the antibody titres we considered assumed the vaccine modified the individuals erythrocyte invasion rate, such that, $\tilde{\beta}_i \sim \mathcal{N}(\mu(1+\delta_{vacc}), \sigma)$, where $\mu$ is the mean log attack rate, $\sigma$ is the standard deviation of the log attack rate, and $\delta_{vacc}$ is the shift in the mean caused by the effect of the vaccine. For the vaccine to have a positive effect on the population we would expect the median of the posterior for $\delta_{vacc}$ to be negative, and for this effect to be significant we would expect the majority of the density of the posterior for $\delta_{vacc}$ to be negative.

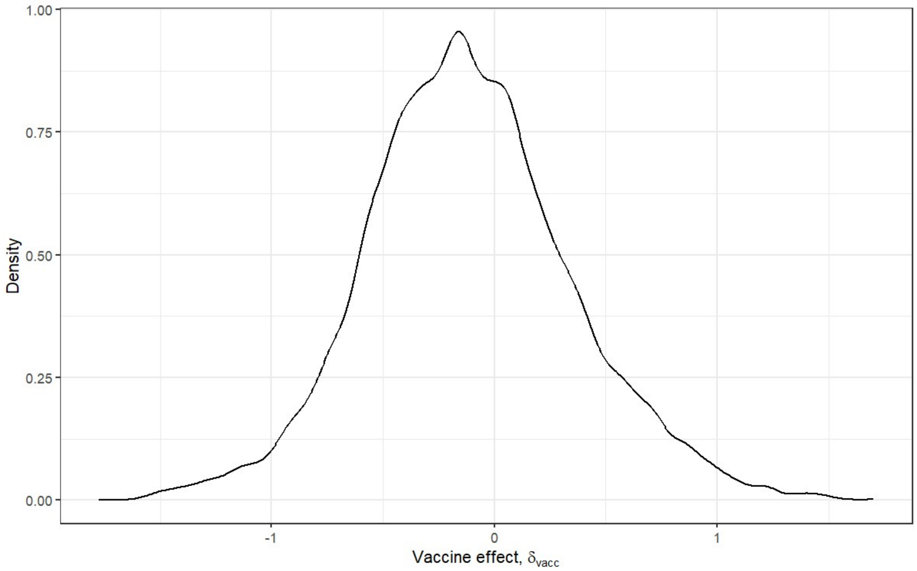


*Figure S1: Posterior density for $\delta_{vacc}$, estimated using the phase 1b trial data for AMA1*

As can be seen in Figure S1, there is a very slight effect of the vaccine – the peak of the posterior is slightly below 0, but this effect is very weak and the domain of support of the posterior comfortably straddles both 0 and positive values. This suggests, as was reported by Payne *et al.*[1] that the effect of the vaccine is negligible.

### Generation of synthetic cohorts

To generate synthetic cohorts, we draw samples from the distributions estimated using the CHMI trial for RH5.1. When recovering the estimates for the reduction in PMR we explicitly draw antibody titres using the estimated distributions. However, when estimating the effects of the vaccine against onwards transmission or the probability of exceeding a parasitaemia threshold we do not explicitly use antibodies. The work investigating those definitions of vaccine efficacy is more focussed on the consequences of a vaccine which is a certain level of efficacy on average. Hence the effect of individual variation caused by antibody titre distributions is not an important consideration.

When recovering the estimates for the reduction in PMR, we sample 12 individuals as in the original trial and allocate them to each arm simultaneously. To do this we draw a $\beta$ value for being in each of the control groups (Groups 6, 8 and 9), subject to the distributions described by the estimated parameters. To simulate the effect of the vaccine we draw two antibody titre values, one corresponding to being within group 5 and one corresponding to being in group 7. We then use these antibody titre values to calculate the reduction in $\beta$ using Equation 5. These 5 $\beta$ values are then used to generate trajectories of parasitaemia over 14 days, before the parameter estimation scheme described in the Methods section is used.

To generate the trajectories used to estimate the vaccine efficacy against onwards transmission and probability of exceeding thresholds we use similar techniques. First in both instances we assume that all vaccinated participants would receive the same benefit from the vaccine. We are more concerned with the public health impacts of a vaccine that is on average a certain level of effective rather than the impacts of antibody titre heterogeneity. We assume that prior to the effect of the vaccine, individuals are distributed as though they belonged to group 6 from the CHMI trial of RH5.1. We allow individual heterogeneity in the development rate and effectiveness of the immune response, with each individual's $\omega$ and $\epsilon$ parameters being log-normally distributed. Both $\omega$ and $\epsilon$ are drawn from the same distribution such that, $\log(\omega) \sim \mathcal{N}(\log(0.001), 0.5)$ and, $\log(\epsilon) \sim \mathcal{N}(\log(0.001), 0.5)$. Taking advantage of the nature of synthetic cohorts, we can simultaneously run each individual as though they both did and did not receive the vaccine. This allows us to both have heterogeneity in the adaptive immune response and be consistent across the two groups. To ensure the effect of the heterogenous immunity is roughly consistent across the explored vaccine efficacies, we simulate 200 individuals in both the vaccinated and control arms.

### Replicating reduction of PMR

To estimate individual PMRs we used a Bayesian scheme, with the same likelihood functions described in Equations 6 and 7 in the Methods. The model described in Equation 1 in the Methods is replaced by a PMR based model described in Equation 1 below. Where $P_0$ is the initial density of parasitaemia, $PMR_i$ is the individual's parasite multiplication rate over two days, and $t$ is the time since challenge in days. Individual PMRs are assumed to be normally distributed, with the vaccine shifting the mean of the distribution. Individuals PMRs are distributed as $PMR_i \sim \mathcal{N}\left((\widehat{\mu_k} + \widehat{\delta_i})(1 - V), \widehat{\sigma_k}\right)$, where $\widehat{\mu_k}$ is the mean PMR for trial round k, $\widehat{\delta_i}$ denotes the effect of one previous challenge, $\widehat{\sigma_k}$ is the standard deviation of the PMRs in trial round k, and $V$ is the effect of the vaccine. There is a relationship between the efficacy observed against PMR and the efficacy observed against erythrocyte invasion rates. However, it depends on the individual erythrocyte invasion rate and individual PMRs and therefore cannot be used to map population level estimates of these efficacies easily (Supplementary Material).

$$\log_{10} P(t) = \log_{10} P_0 + \frac{t}{2}\log_{10} PMR \qquad (1)$$

Here we are estimating $P(\boldsymbol{PMR}, \boldsymbol{\mu}, \boldsymbol{\sigma}, \boldsymbol{\delta}, V|\boldsymbol{x})$, where $\boldsymbol{x}$ is the individual level parasitaemia data.

We then simulate several trajectories using the estimated parameters and the model described in Equation 1 (Methods section). Using these simulated trajectories, we estimate the PMR values and the effect of the vaccine in the same way as using the trial data. This allows us to have another form of validation, wherein we can both replicate the results from Minassian *et al.* (2019)[2] and show that our model produces similar estimates.

**Recovering estimates of PMR reduction**

Using the model described in Equation 1, we recover estimates for the effect of the vaccine and the mean PMRs that are highly comparable to those in the phase I/IIa RH5 trial[2]. For the control group in the first round of the trial, we estimate a mean PMR of 9.05 (95% CI, 7.6, 10.3) with standard deviation 2.44 (95% CI, 1.75, 3.91). Similarly for the second round of the trial, we estimate a mean PMR of 13.0 (95% CI, 10.9, 15.0) with standard deviation 2.22 (95% CI, 1.47, 3.71). Our estimate for the effect of the previous challenge on the rechallenged individuals is a change in the mean PMR of -1.02 (95% CI, -3.77, 1.72), suggesting the previous challenge had a limited effect on the PMR of the individuals. Finally, our estimate for the effect of the vaccine is a proportional reduction in the mean of the PMR by 23.5% (95% CI, 14.1%, 33.2%). In all instances here the median of our posterior estimates fall within the confidence intervals of the estimates for these parameters from Minassian *et al.* (2019). Crucially, using a small modification (see Methods section) of the parameter estimation technique used to fit the asexual blood-stage model described in Equation 1 we have recovered the results of Minassian *et al.* (2019), lending credence to our estimated parameters.

We simulated a synthetic cohort of trial participants whose antibody titre levels and parasite erythrocyte invasion rates were generated using the distributions estimated for the phase 1 trial (see Supplementary Material). We then recalculated the effects of the vaccine on PMR, replicating the analysis described above using the synthetic cohort. The results from the synthetic cohort are broadly comparable to the results we estimated for the phase 1 trial. The effect of the vaccine is a proportional reduction in PMR of 21.7% (95% CI, 14.0%, 29.9%), almost identical to the vaccine efficacy estimated using the trial data. From the control group in the first round, we estimate a mean PMR of 10.2 (95% CI, 8.19, 11.9) with standard deviation 3.13 (95% CI, 2.1, 5.16). For the control group in the second round, we estimate a mean PMR of 12.0 (95% CI, 11.1, 12.9) with standard deviation 1.67 (95% CI, 1.27, 2.33). Our estimate of the effect of the previous challenge is a change in the mean PMR of -1.62 (95% CI, -2.94, -0.30). The estimated parameters for the PMR model are slightly different than the estimates for the trial data, but in almost all instances the median of the trial estimate is within the credible interval for the estimates of the simulated data.

### **Including outliers in vaccine parameter estimation**

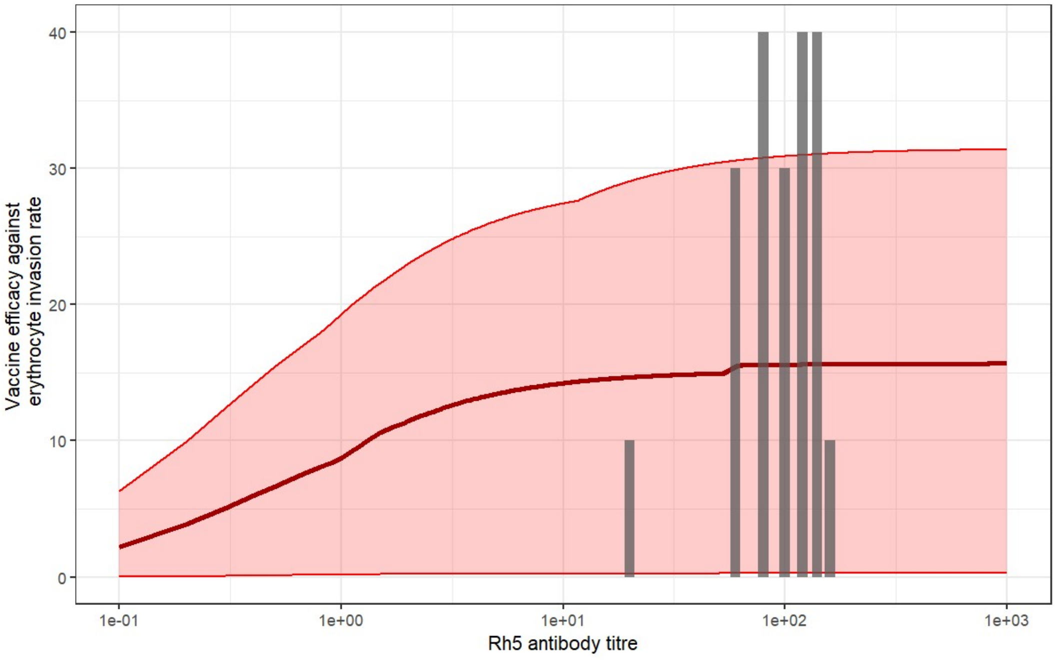


*Figure S2: Relationship between vaccine efficacy against erythrocyte invasion rate and RH5 antibody titre (µg/mL). The red line shows the median relationship between antibody titre and vaccine response, and the shaded region shows the 95% credible interval. Vertical grey bars show the distribution of the antibody titres from the challenge study participants.*

As can be seen in Figure S2, the inclusion of the outliers reduces the efficacy of the vaccine, but the confidence interval for the vaccine still demonstrates a positive effect. The two outlier individuals were both removed from the rechallenge control group, as 1 had a lower PMR when challenged the second time compared to when they were challenged the third time, and the other had no parasite growth over the whole challenge.

### Sensitivity Analysis

Here we vary the parameters shown in Table 2 in the main text and replicate the plots shown in Figure 2 using the posterior estimates generated using the altered parameter values with the same method as in the main text. We observe that whilst the plots have slight numerical differences in the estimated values, the qualitative behaviour and shapes of the plots are the same.

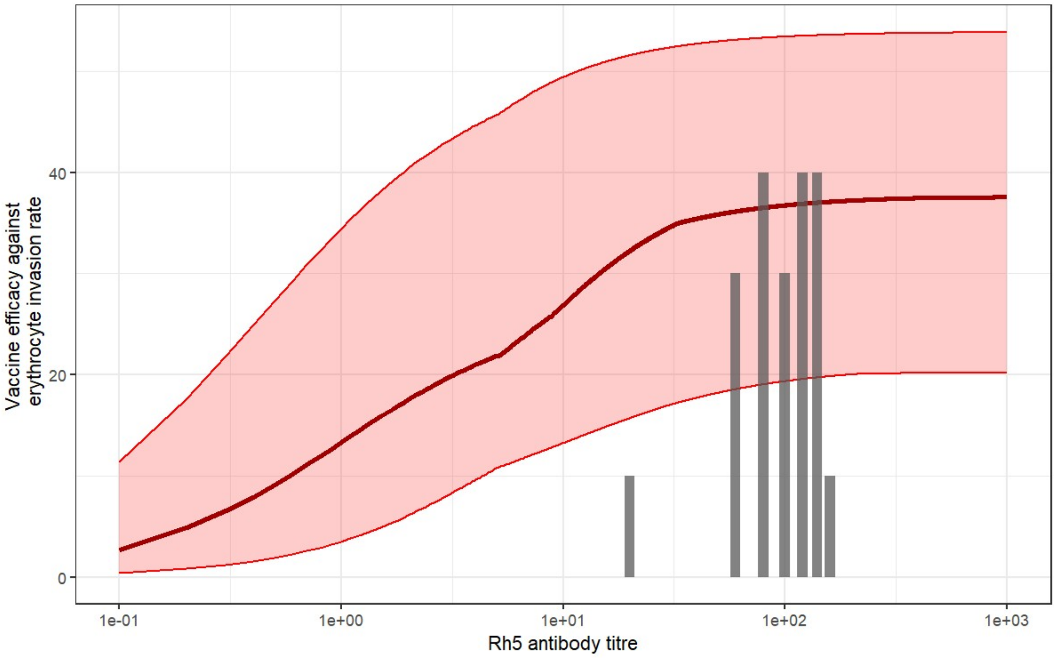


*Figure S3: Relationship between vaccine efficacy against erythrocyte invasion rate and RH5 antibody titre ($\mu g/mL$). The red line shows the median relationship between antibody titre and vaccine response, and the shaded region shows the 95% credible interval. Vertical grey bars show the distribution of the antibody titres from the challenge study participants. Parameters as in Table 1 except $d = 0.0$..*

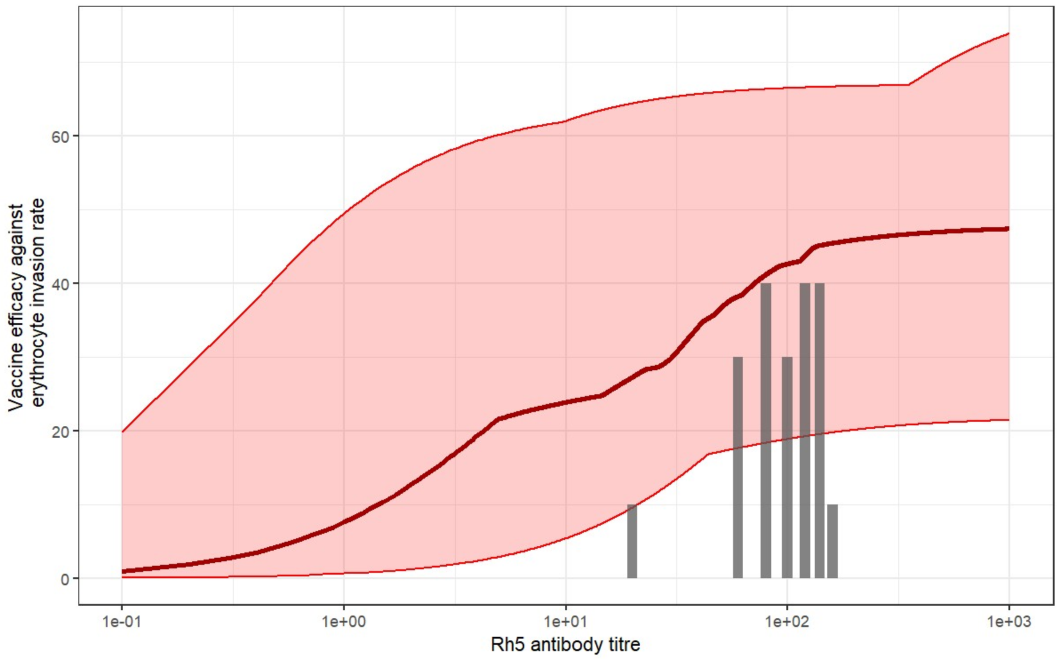


*Figure S4: Relationship between vaccine efficacy against erythrocyte invasion rate and RH5 antibody titre* ($\mu g/mL$). *The red line shows the median relationship between antibody titre and vaccine response, and the shaded region shows the 95% credible interval. Vertical grey bars show the distribution of the antibody titres from the challenge study participants. Parameters as in Table 1 except* $d = 0.0, r = 16$

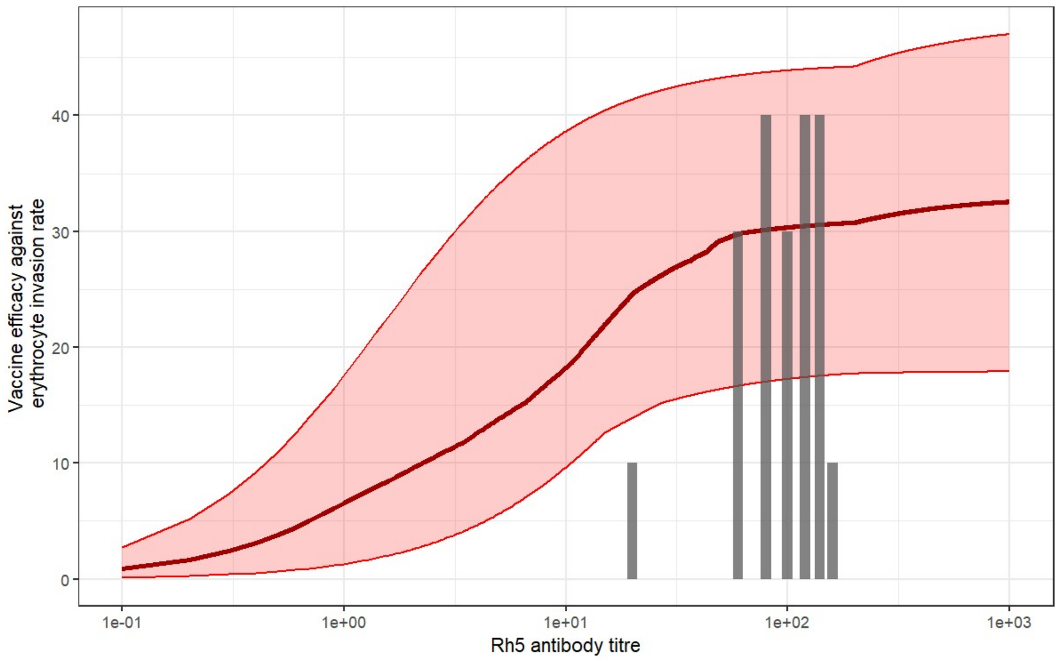


*Figure S5: Relationship between vaccine efficacy against erythrocyte invasion rate and RH5 antibody titre* ($\mu g/mL$). *The red line shows the median relationship between antibody titre and vaccine response, and the shaded region shows the 95% credible interval. Vertical grey bars show the distribution of the antibody titres from the challenge study participants. Parameters as in Table 1 except* $d = 10$

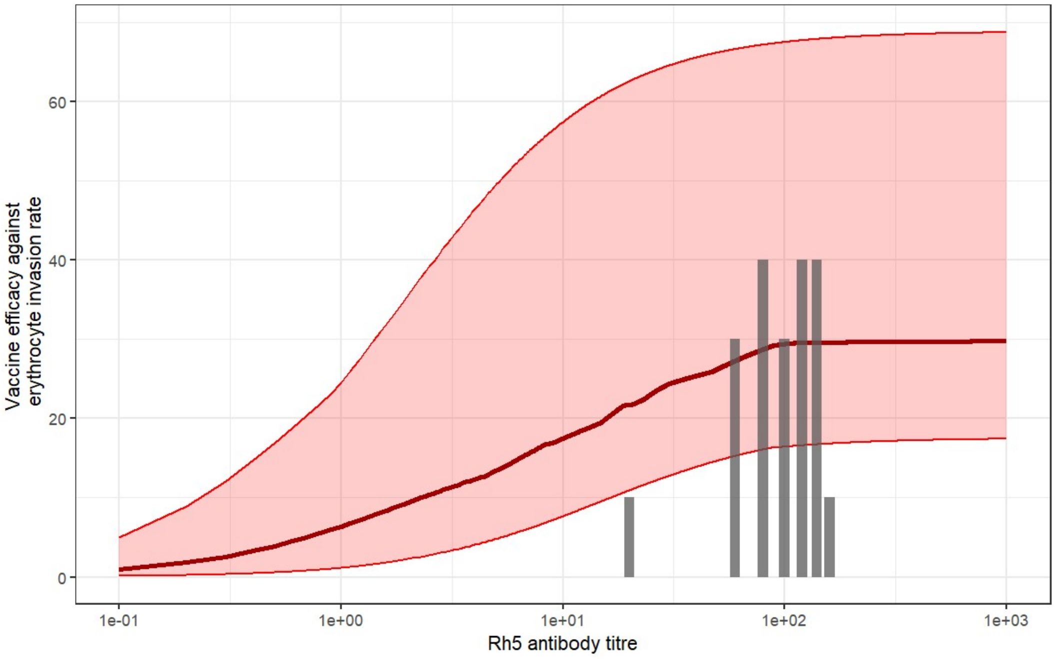


*Figure S6: Relationship between vaccine efficacy against erythrocyte invasion rate and RH5 antibody titre* ($\mu g/mL$)*. The red line shows the median relationship between antibody titre and vaccine response, and the shaded region shows the 95% credible interval. Vertical grey bars show the distribution of the antibody titres from the challenge study participants. Parameters as in Table 1 except* $\sigma_{obs} = 0.25$

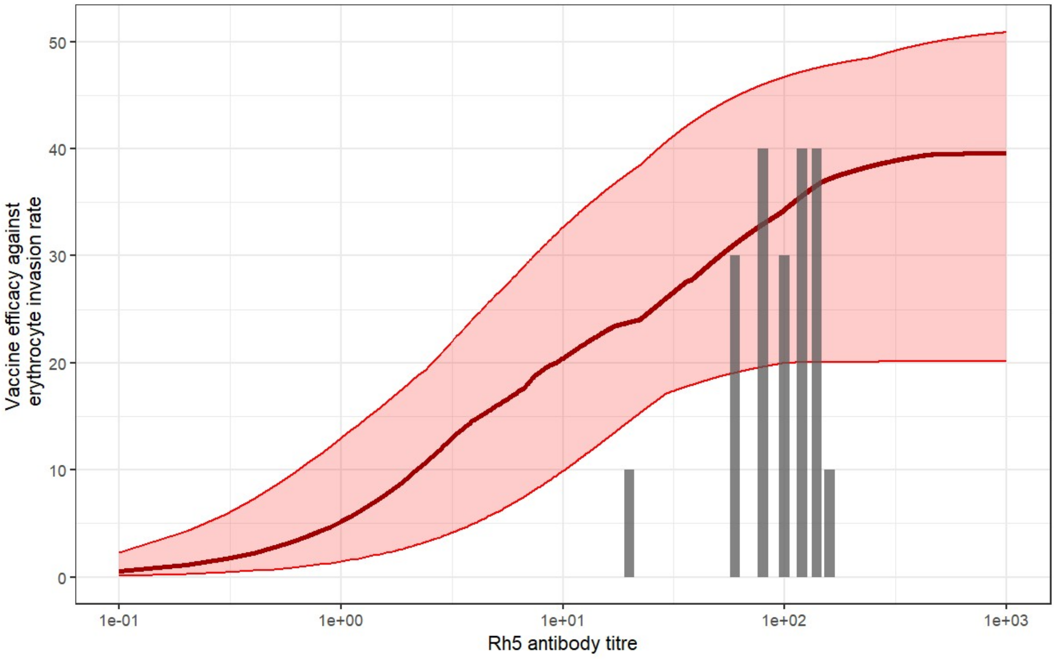


*Figure S7: Relationship between vaccine efficacy against erythrocyte invasion rate and RH5 antibody titre* ($\mu g/mL$)*. The red line shows the median relationship between antibody titre and vaccine response, and the shaded region shows the 95% credible interval. Vertical grey bars show the distribution of the antibody titres from the challenge study participants. Parameters as in Table 1 except* $\sigma_{obs} = 1$

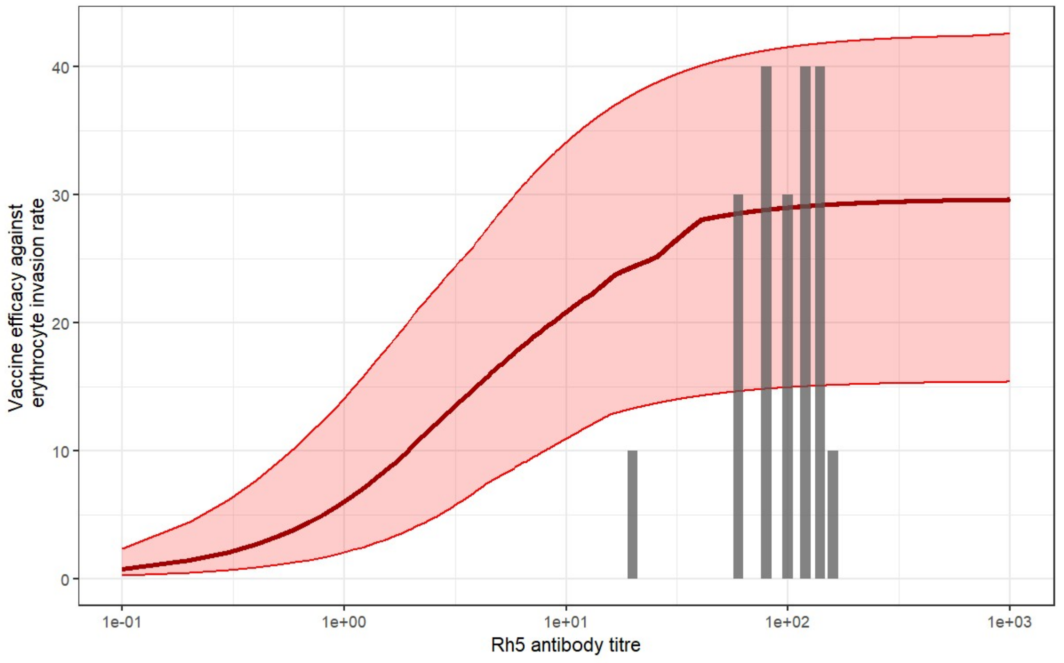


*Figure S8: Relationship between vaccine efficacy against erythrocyte invasion rate and RH5 antibody titre* ($\boldsymbol{\mu g/mL}$)*. The red line shows the median relationship between antibody titre and vaccine response, and the shaded region shows the 95% credible interval. Vertical grey bars show the distribution of the antibody titres from the challenge study participants. Parameters as in Table 1 except* $r = 35$

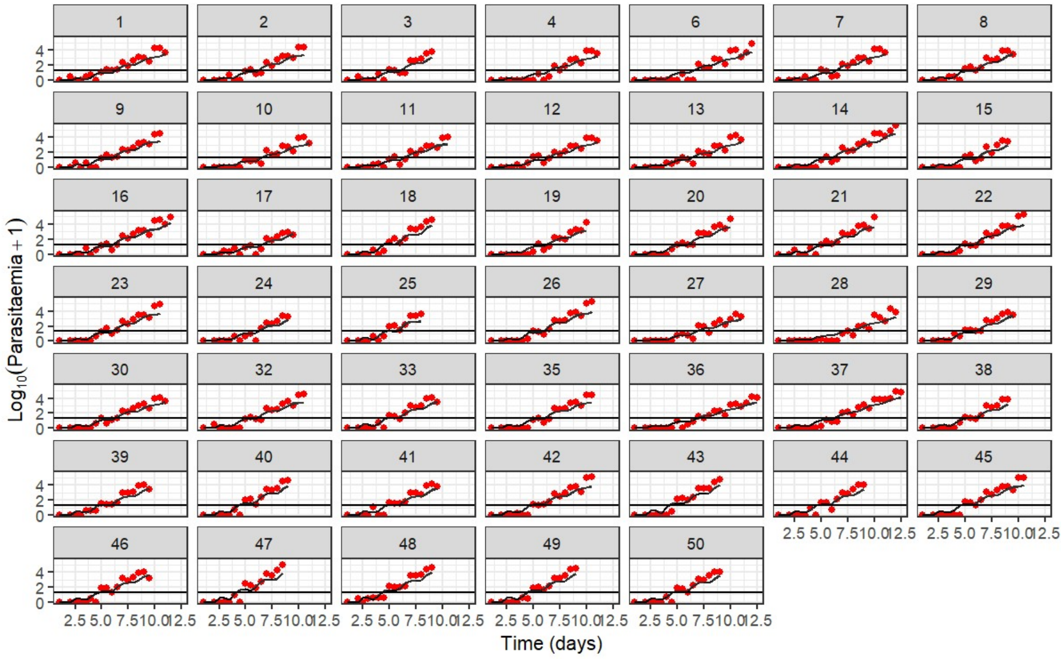


*Figure S95: Individual fits to parasitaemia data. Red dots denote the actual observed values for that individual, black lines shows median model estimate for that individual and the shaded regions represent uncertainty estimates.*

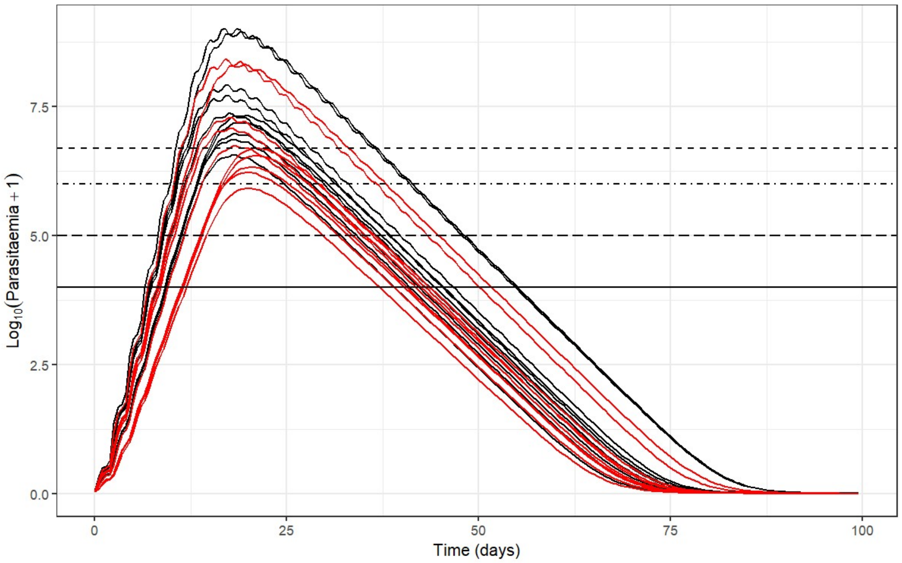


*Figure S10: 10 realisations of the dynamics described by Equation 2 (Methods). The red lines show the dynamics of the vaccinated individuals and the black lines show the dynamics of unvaccinated individuals.*
